# Pilot Plant-Based Lifestyle Medicine Program in an Urban Public Healthcare System: Evaluating Demand and Implementation

**DOI:** 10.1101/2022.02.09.22270738

**Authors:** Stephanie L. Albert, Rachel Massar, Lorraine Kwok, Lily Correa, Krisann Polito-Moller, Shivam Joshi, Sapana Shah, Michelle McMacken

## Abstract

**Background:** Lifestyle interventions that optimize nutrition, physical activity, sleep health, social connections, and stress management, and address substance use can reduce cardiometabolic risk. Despite substantial evidence that healthful plant-based diets are beneficial for long-term cardiometabolic health and longevity, uncertainty lies in how to implement plant-based lifestyle programs in traditional clinical settings, especially in safety-net contexts with finite resources.

**Methods:** In this mixed-methods implementation evaluation of the Plant-Based Lifestyle Medicine Program piloted in a large public healthcare system, we surveyed participants and conducted qualitative interviews and focus groups with stakeholders to assess program demand in the eligible population, and feasibility of implementation within the safety-net setting.

**Findings:** Program demand was high and exceeded capacity. Participants’ main motivations for joining the program included gaining more control over life, reducing medication, and losing weight. The program team, approach, and resources were successful facilitators. However, the program faced administrative and payor-related challenges within the safety-net setting, and participants reported barriers to access.

**Conclusions:** Stakeholders found the program to be valuable, despite challenges in program delivery and access. Findings provide guidance for replication. Future research should focus on randomized controlled trials to assess clinical outcomes as a result of program participation.

## INTRODUCTION

Lifestyle behaviors play a critical role in the risk of chronic disease and premature death. Up to 80% of premature deaths could be prevented by adhering to a healthful diet, being physically active, and not smoking.[1] Moreover, in the United States, poor-quality diets are the leading risk factor for dying of a chronic disease.[2] Thus, interventions that support positive lifestyle changes have great potential to reduce the burden of chronic disease and premature mortality.

Research has shown that intensive lifestyle change programs reduce angina and cardiac events, and may slow progression of atherosclerotic plaques, while improving cardiometabolic risk profiles by lowering weight, blood pressure, fasting lipids, and blood sugar.[3, 4] The lifestyle interventions are centered around optimizing nutrition, physical activity, sleep, and social connections while managing stress and avoiding harmful substance use. Regarding nutrition, research from multiple lines of evidence suggests that an optimal diet for long-term cardiometabolic health and longevity is a healthful plant-based diet -- that is, an eating pattern that emphasizes a large proportion of whole or minimally processed plant foods (vegetables, whole fruits, legumes, whole grains, nuts, and seeds) while reducing red meats, processed meats, refined grains, and other highly processed foods, particularly those high in sodium, added sugars, and saturated fats.[5-11] In addition, published evidence demonstrates the benefits of a healthful plant-based diet for the treatment of chronic conditions such as type 2 diabetes, hyperlipidemia, obesity, hypertension, and atherosclerotic heart disease.[3, 12-16]

Lifestyle medicine is defined by the American College of Lifestyle Medicine as the use of evidence-based, lifestyle therapeutic interventions—including a whole-food, plant-predominant eating pattern, regular physical activity, restorative sleep, stress management, avoidance of risky substances, and positive social connection—as a primary modality, delivered by clinicians trained in these modalities, to prevent, treat, and often reverse disease.[17] Historically, many plant-based lifestyle medicine programs have operated outside of the traditional healthcare system, offered in community-based programs, “jumpstarts”, or residential programs with participants paying out of pocket; as part of worksite or insurance wellness programs; or in research studies.[3, 4, 18, 19] In recent years, increasing numbers of traditional healthcare systems are adopting lifestyle medicine programs.[20] Safety-net hospitals provide care to underserved communities and other populations that face a disproportionately high burden of chronic diseases, and thus could stand to benefit the most from participation in a lifestyle medicine program.

However, uncertainty lies in how to implement plant-based lifestyle medicine programs in traditional clinical settings, especially in the safety-net context with finite resources. Moreover, research is limited on optimal ways to design and implement plant-based lifestyle medicine programs among culturally diverse patient populations, including individuals facing significant environmental and socioeconomic barriers to making lifestyle changes.

The Plant-Based Lifestyle Medicine (PBLM) Program is, to our knowledge, the first lifestyle medicine program of its kind to be piloted within a traditional, safety-net healthcare setting. We conducted a mixed-methods implementation evaluation using survey data and qualitative interviews and focus groups to assess the demand for the program in the eligible participant population, and feasibility of implementation within the safety-net healthcare context. Our study provides valuable lessons learned for others seeking to implement lifestyle medicine programs within a traditional healthcare setting.

## INTERVENTION

The PBLM Program was implemented as a one-year pilot program within an adult primary care center in the NYC Health + Hospitals (NYC H+H) system, the largest public healthcare system in the United States. The program’s central goal was to help participants reduce their cardiometabolic risk through positive lifestyle changes, including a healthful plant-based diet, physical activity, sleep, stress reduction, social connection, and avoidance of risky substances. The pilot PBLM Program received one year of funding from NYC H+H to serve between 100-200 participants.

Program leadership sought to construct a team that would have a broad range of knowledge and experience to support participants’ lifestyle modifications. The team included four physicians who had varying areas of specialty or interest (i.e., internal medicine, cardiology, nephrology, and musculoskeletal disorders), one registered dietitian, and one health coach. All team members followed a plant-based diet. Additionally, the registered dietitian, a trained chef, had training and experience in public health and community-focused nutrition education. The health coach had certifications in yoga therapy, personal fitness training, and healthful plant-based nutrition as well as experience with programs to improve stress management and sleep behaviors. The program dietitian and two of the physicians were proficient or fluent in Spanish.

The eligibility criteria for participation in the PBLM Program included adults with prediabetes, type 2 diabetes, hypertension, heart disease, dyslipidemia, and/or excess weight (BMI ≥25). Participant recruitment consisted of various types of media outreach (social media, broadcast news, podcast interviews, newsletters/newspapers, and announcements from the office of the Brooklyn Borough President), as well as very limited physician referrals from within the NYC H+H location. Interested participants were directed to call a dedicated contact center team to join a waitlist. Eligible participants were enrolled from the waitlist on a first come first serve order.

The initial entry to the program involved a visit with a program physician for an individualized, comprehensive evaluation of the participant’s goals, comorbidities, medications, family/cultural/social context, current lifestyle habits, and readiness and barriers to change. They also provided preliminary education on the benefits of a healthful plant-based diet for reducing chronic disease risks, made recommendations around diet and other lifestyle changes, and developed an individualized plan for monitoring and follow-up. Physicians monitored participants’ clinical outcomes and adjusted their medications as needed throughout their participation in the program, seeing each participant at least every 2-3 months, and more frequently when needed.

Participants then had separate individual consults with the registered dietitian and health coach. The registered dietitian performed in-depth dietary assessments and Medical Nutrition Therapy tailored to participants’ needs and worked with participants to create action plans that were focused on diet as well as other lifestyle behaviors. The health coach provided support to participants in implementing the nutrition plan recommended by the dietitian, troubleshooting common issues such as meal planning. Additionally, the health coach focused on helping participants to make other healthful lifestyle behavior changes including physical activity, sleep, and stress management, with an emphasis on overall well-being. After the initial consultations, participants met with the registered dietitian and health coach alternating every two weeks, as schedules allowed, to monitor progress and set new action plans. Physician, dietitian, and health coach recommendations were tailored to each participant’s specific socioeconomic and cultural background, taking into account access to healthy foods, the neighborhood-built environment, family situation, and other key factors.

Resources available to all participants included a healthful plant-based diet starter guide developed by program staff, plant-based cookbook(s), a Healthy Savings card [21] for grocery store discounts on fresh produce, Health Bucks [22] ($2 coupons that can be used to purchase fresh fruits and vegetables at all NYC farmer’s markets), and a private Facebook group where participants could share ideas and provide support to each other.

Group visits were added to the program a few months after the initial program launch to increase participant engagement and peer support while utilizing limited resources (time and personnel) most efficiently. The registered dietitian and health coach led two group classes per week, teaching an 8-session rolling curriculum covering the basics of plant-based nutrition, mindfulness, stress reduction, navigating social situations, recipe conversions, meal preparation, reading food labels, and physical activity. In addition, an exercise trainer offered classes focused on aerobic and strength training, using resistance bands that the program was able to provide to patients.

The program was designed to fit largely within the existing finance structure of the hospital system, with physician and dietitian visits being billed to insurance as per hospital protocols. Health coach services were funded by the system and not billable. The finance workflow plan included reaching out to patients in advance of their initial visit to review insurance status and coverage of physician services. Patients who lacked insurance were billed according to a sliding fee scale as would occur for any other hospital visit. The program was supported administratively by a coordinator working several hours a week. During the pilot phase, participants used existing methods for contacting the care team, the same ways accessible to non-program patients receiving care in the ambulatory care setting. Of note, three months into the pilot phase, the hospital system transitioned to a new electronic health system, which involved a significant learning curve for all staff members (providers, finance, administration).

This description of the PBLM Program reflects its design at the outset of the pilot phase. As anticipated, continuous assessments and adjustments were made throughout the pilot period to adapt to the realities of working within a complex healthcare setting, as well as to meet the evolving needs of the participants.

## METHODS

The primary goals for the pilot PBLM Program evaluation were to assess demand for the program in the eligible participant population and the feasibility of implementing the program within the safety-net healthcare setting. The NYU Grossman School of Medicine (NYUGSOM) evaluation team used a mixed-methods approach to collect both quantitative and qualitative data to capture the perspectives of different groups of key program stakeholders (program participants and providers). Approval for this study was obtained from both the NYUGSOM IRB as well as the Office of Research and Administration for Implementation at NYC Health + Hospitals/Bellevue.

### Quantitative Data

#### Survey Data

Data come from surveys of program participants at two time points (baseline, 6-month). All individuals who signed up for an initial visit in the PBLM Program were invited to participate in the evaluation (n=173). Of those, 131 agreed to be contacted by the NYU evaluation team and 111 provided verbal consent (85%). Subsequently, 109 PBLM Program participants completed a baseline survey (98%), and 84 individuals completed a 6-month survey (76%). Baseline surveying began on January 29, 2019 and continued until July 30, 2019, and the 6-month data were collected between July 30, 2019 and February 26, 2020. Surveys were administered telephonically by trained evaluation staff and took approximately 25 minutes to complete. Respondents were given a $10 gift card each time a survey was completed to thank them for their time and effort.

#### Measures

##### Participant Characteristics

The survey included questions on demographic characteristics including gender, age, race and ethnicity, language use, education, health insurance, marital status, and food security. To assess experience with weight stigma, participants were asked if they agreed or disagreed with the statement, “People have judged you because of your weight” on a 4-point Likert scale. To assess neighborhood food environment, we adopted three questions used by Ortega et al. [23] Participants were asked to rate their agreement with statements about the availability, affordability, and quality of healthy foods in their neighborhood on a 4-point Likert scale. To assess self-rated health, we utilized one question from the Lifestyle Assessment Form (long version) created by the American College of Lifestyle Medicine & Loma Linda University Health [24], where participants rated their level of health on a scale from 0 to 10, with 0 being “very poor health” and 10 being “excellent health.” Quality of life in the past month was measured using a question from the World Health Organization Quality of Life-BREF measure [25] using a 5-point Likert scale that ranged from “very poor” to “very good.”

##### Motivation and Confidence

To assess overall motivation and confidence in adopting and maintaining a plant-based diet, we adapted two questions from Lee at al.[26] Participants were asked to rate two statements: “You are motivated to eat a plant-based diet because you want to be healthy” and “You are confident in your ability to stick to a plant-based diet to be healthy” using a 4-point Likert scale from “not at all true” to “very much true.” We also adapted one question from the Treatment Motivation Questionnaire [27] to measure participants’ level of agreement with the statement: “A plant-based diet will work for you” using a 4-point Likert scale. Additionally, participants were asked a series of questions to identify one health condition and one lifestyle behavior they were most motivated to change, as well as rate the level of importance and confidence in making these changes on a scale from 0-10, with 0 being “not at all important/confident” and 10 being “very important/confident.” [24]

##### Ease of Participation and Program Satisfaction

We measured ease of participation and program satisfaction during the 6-month survey administration with questions developed *de novo* by the research team. Participants rated the level of ease or difficulty of various administrative aspects of participating in the program, such as scheduling appointments, getting insurance and billing questions answered, and communicating with program and clinic staff using a 4-point Likert scale from “very difficult” to “very easy.” To measure program satisfaction, participants rated their level of agreement with statements about various aspects of the program related to the program providers and program content using a 4-point Likert scale from “strongly disagree” to “strongly agree.” For example, “The information you were given about plant-eating patterns was easy to understand.”

##### Analyses

We present descriptive statistics for all survey measures. Participant characteristics and motivation and confidence are presented from the baseline survey administration to describe the sample at the start of program participation. Data on ease of participation and program satisfaction are presented from the 6-month survey administration, in order to report on participants’ experience after spending adequate time in the program.

### Qualitative Data

Qualitative data come from key informant interviews and focus groups. All interviews and focus groups were conducted by two experienced interviewers. One interviewer acted as the primary interviewer while the second interviewer took notes and asked clarifying questions. All interviews were recorded and professionally transcribed.

#### Key Informant Interviews

The evaluation team conducted key informant interviews with all 6 program providers. Interviews with program providers included topics such as recruitment, facilitators and barriers to implementation, suggestions for improvement, and sustainability. The interviews were conducted in person between June and July 2019 and lasted between 45-60 minutes. No incentives were provided to program providers for participation.

Interviews were also conducted with 5 participants who had dropped out of the program. Interview questions covered topics including reasons for participation, reasons for leaving the program, experience during participation, and suggestions for improvement. Additional interviews were planned but saturation was quickly reached and no additional themes emerged after the first few interviews. Interviews were conducted over the phone between August and December 2019. Interviews lasted between 10-20 minutes and participants received a $10 gift card for their participation.

#### Focus Groups

The team conducted two focus groups with program participants (n=17). In order to be eligible, participants had to agree to be contacted about the focus groups at the time of consent. The evaluation team partnered with the PBLM registered dietitian and health coach to develop a prioritized list of eligible focus group participants such that it maximized diversity in age, gender, time in the program, and success within the program. Focus group questions covered topics including reasons for participation, facilitators and barriers of the program, benefits and challenges to participation, outcomes of participation, suggestions for improvement, and sustainability of participation. The focus groups were conducted in-person at NYUGSOM offices between October and November 2019. Each focus group lasted approximately 90 minutes and participants received a $25 gift card for participating.

#### Analyses

The evaluation team members who conducted the interviews and focus groups met to discuss emerging themes, monitor thematic saturation, and develop *a priori* coding scheme (deductive approach) closely aligned with the interview and focus group protocols. To establish consistency between coders, two trained members of the research team individually coded two key informant interviews and one focus group transcript using ATLAS.ti software and met to compare coding decisions and resolve discrepancies. Once agreement was reached between coders, minor changes were made to the coding scheme and the remaining interview and focus group transcripts were divided between the two coders. All transcripts were coded using the final schema.

## FINDINGS

Findings are grouped into four categories: 1) participant characteristics; 2) demand and participant motivation; 3) implementation facilitators and challenges; and 4) program satisfaction. When available, we present both quantitative and qualitative data and include the perspectives of the multiple stakeholder groups on each topic.

### Program Participant Characteristics

Table 1 shows the characteristics of survey participants at baseline. The sample was majority female, participants were on average a little older than 50 years old, about one-third of the sample identified as Black/African American and one-third identified as White, and the vast majority spoke only English at home. The majority reported their highest educational attainment was college or more than college, over half of the sample indicated that they had private insurance, and the majority had high food security. More than half of the sample had experienced weight-related stigma. The majority of participants agreed that they had access to high quality foods in their neighborhoods. On average participants rated their own health as 5.9 out of 10, and just over half described their quality of life as being “good” or “very good.”

**Table 1:** Characteristics of the PBLM Program Participants (n=109)

|  | **N** | **Percent or Mean (SD)** |
| --- | --- | --- |
| **Gender** | 106 |  |
| Male |  | 33.0 |
| Female |  | 67.0 |
| **Age** | 106 | 53.0 (12.0) |
| **Race and ethnicity** | 105 |  |
| American Indian or Alaska Native |  | 1.0 |
| Asian |  | 5.7 |
| Black or African American |  | 39.0 |
| Hispanic or Latino |  | 17.1 |
| Native Hawaiian or Other Pacific Islander |  | 0.0 |
| White |  | 31.4 |
| Two or more |  | 2.9 |
| Other |  | 2.9 |
| **Language use** | 107 |  |
| English-only |  | 85.0 |
| Spanish-only |  | 5.6 |
| English and Spanish |  | 4.7 |
| Other language |  | 4.7 |
| **Education** | 107 |  |
| Less than high school |  | 4.7 |
| High school |  | 6.5 |
| Some college |  | 26.2 |
| College/More than college |  | 62.6 |
| **Health insurance** | 106 |  |
| Private (employer, else's employer, purchased) |  | 59.4 |
| Public (Medicaid, Medicare) |  | 21.7 |
| No insurance |  | 1.9 |
| Other (other, two or more, military, COBRA) |  | 18.4 |
| **Marital status** | 107 |  |
| Single |  | 43.0 |
| Married/living with partner |  | 43.9 |
| Separated/divorced/widowed |  | 13.1 |
| **Food security** | 109 |  |
| Often we don't have enough to eat |  | 0.0 |
| Sometimes we don't have enough to eat |  | 2.8 |
| We have enough to eat but not always the kinds of food we want |  | 43.1 |
| We always have enough to eat and the kinds of food we want |  | 54.1 |
| **Weight Stigma** | 105 |  |
| People have judged you because of your weight (agree/strongly agree) |  | 56.2 |
| **Neighborhood Food Environment (agree/strongly agree)** |  |  |
| Healthy foods sold in your neighborhood are too expensive | 106 | 66.0 |
| There are places in your neighborhood where you can buy healthy foods | 107 | 86.9 |
| The healthy foods sold in your neighborhood are low quality | 104 | 33.7 |
| **Self-rated health (0-10)** | 107 | 5.9 (1.8) |
| **Quality of life (good/very good)** | 107 | 53.3 |

NOTES: Ns may vary due to missing data.

### Program Demand and Participant Motivation

The PBLM Program generated an enormous amount of interest with more than 850 individuals adding their name to a waitlist. The vast majority of program participants were self-referred to the program. Multiple program participants reported first hearing about the program through social media platforms primarily because they were already interested in plant-based nutrition and/or were familiar with the Program Director. Other participants reported seeing or reading about the program on the news or in the local newspaper. A PBLM provider similarly reported that media recruitment was the most successful way of bringing in program participants. Physician referrals to the program was expected to be a major recruitment source; however, the high number of self-referred participants quickly filled up the program waitlist, and did not allow many physician-referred participants to join the program. A PBLM provider also explained that other physicians were not well informed about the program or plant-based nutrition in order to provide referrals to the program.

Table 2 shows that, at baseline, survey participants reported they were highly motivated to adopt a healthful plant-based diet to improve their health, were confident in their ability to maintain a healthful plant-based diet to be healthy, and had an almost universal belief that a healthful plant-based diet would work for them.

**Table 2:**
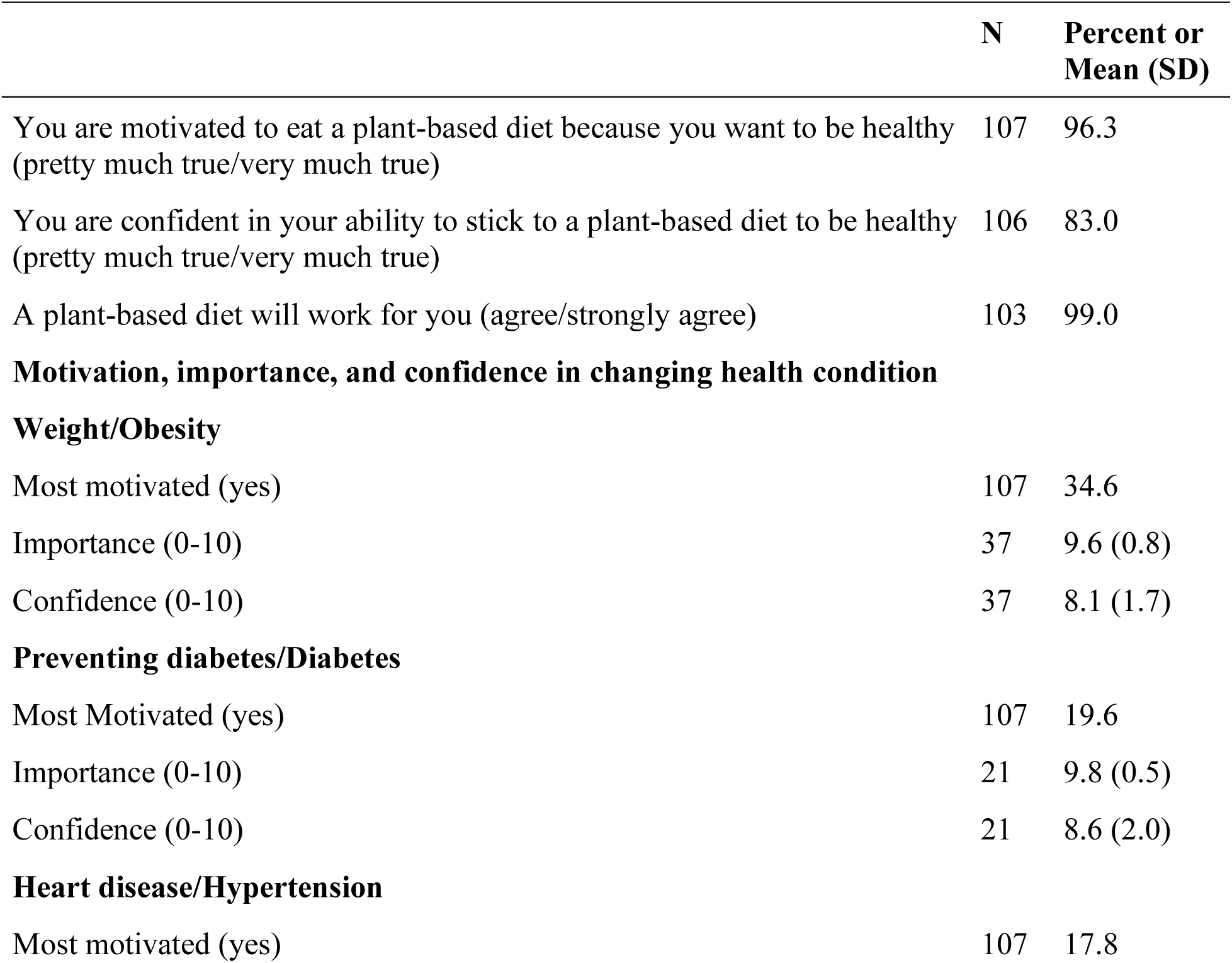
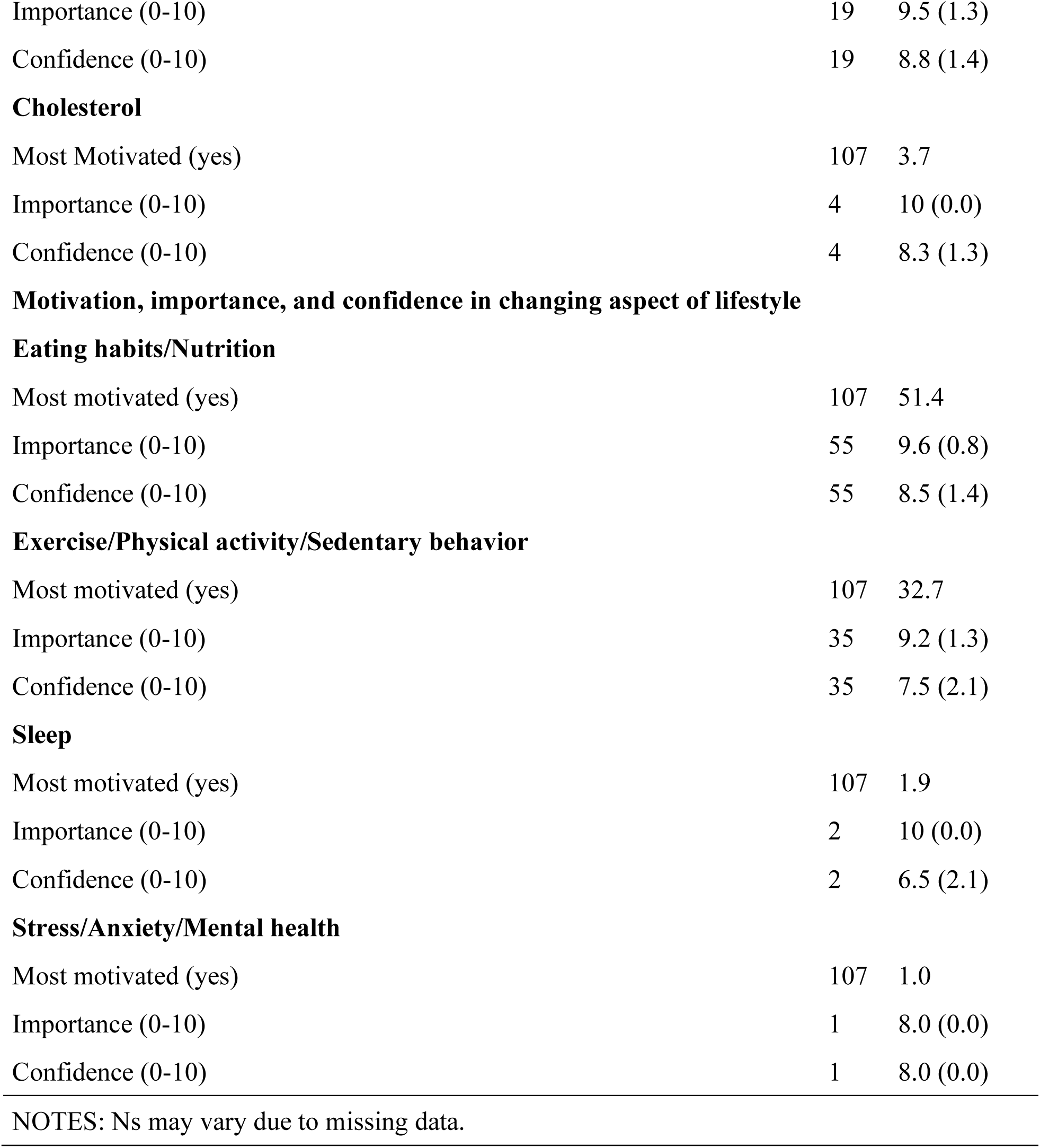
Motivation and Confidence at Baseline (n=109)

|  | **N** | **Percent or Mean (SD)** |
| --- | --- | --- |
| You are motivated to eat a plant-based diet because you want to be healthy (pretty much true/very much true) | 107 | 96.3 |
| You are confident in your ability to stick to a plant-based diet to be healthy (pretty much true/very much true) | 106 | 83.0 |
| A plant-based diet will work for you (agree/strongly agree) | 103 | 99.0 |
| **Motivation, importance, and confidence in changing health condition** |  |  |
| **Weight/Obesity** |  |  |
| Most motivated (yes) | 107 | 34.6 |
| Importance (0-10) | 37 | 9.6 (0.8) |
| Confidence (0-10) | 37 | 8.1 (1.7) |
| **Preventing diabetes/Diabetes** |  |  |
| Most Motivated (yes) | 107 | 19.6 |
| Importance (0-10) | 21 | 9.8 (0.5) |
| Confidence (0-10) | 21 | 8.6 (2.0) |
| **Heart disease/Hypertension** |  |  |
| Most motivated (yes) | 107 | 17.8 |
| Importance (0-10) | 19 | 9.5 (1.3) |
| Confidence (0-10) | 19 | 8.8 (1.4) |
| **Cholesterol** |  |  |
| Most Motivated (yes) | 107 | 3.7 |
| Importance (0-10) | 4 | 10 (0.0) |
| Confidence (0-10) | 4 | 8.3 (1.3) |
| **Motivation, importance, and confidence in changing aspect of lifestyle** |  |  |
| **Eating habits/Nutrition** |  |  |
| Most motivated (yes) | 107 | 51.4 |
| Importance (0-10) | 55 | 9.6 (0.8) |
| Confidence (0-10) | 55 | 8.5 (1.4) |
| **Exercise/Physical activity/Sedentary behavior** |  |  |
| Most motivated (yes) | 107 | 32.7 |
| Importance (0-10) | 35 | 9.2 (1.3) |
| Confidence (0-10) | 35 | 7.5 (2.1) |
| **Sleep** |  |  |
| Most motivated (yes) | 107 | 1.9 |
| Importance (0-10) | 2 | 10 (0.0) |
| Confidence (0-10) | 2 | 6.5 (2.1) |
| **Stress/Anxiety/Mental health** |  |  |
| Most motivated (yes) | 107 | 1.0 |
| Importance (0-10) | 1 | 8.0 (0.0) |
| Confidence (0-10) | 1 | 8.0 (0.0) |

NOTES: Ns may vary due to missing data.

Roughly one third (34.6%) of participants reported wanting to address overweight and obesity through program participation, followed by preventing diabetes/diabetes (19.6%), heart disease/hypertension (17.8%), and cholesterol (3.7%). When asked about lifestyle behaviors, most participants reported motivation to change their eating habits/nutrition (51.4%) or their exercise/physical activity/sedentary behavior (32.7%), while a smaller number were most motivated to address sleep (1.9%) and stress/anxiety/mental health (1.0%). Participants generally reported high levels of importance (8.0-10 out of 10) and confidence (6.5-8.5 out of 10) in addressing health issues and lifestyle behaviors.

PBLM providers and participants reported reasons for joining the program. Multiple PBLM providers spoke about how lifestyle changes were attractive to program participants as a way of improving their health without additional pharmacotherapy or medical procedures.

> There are a lot of people that want more control over their lives and that don’t want as much dependence on the healthcare system and they’re frustrated with taking a lot of medications, different procedures, or seeing their friends or family undergo procedures. … I think there’s a growing movement…for more lifestyle measures, in general, and prevention, in general. (PBLM Provider)

Similarly, several PBLM participants discussed how they joined the program with the intention of reducing their medication burden by controlling their health conditions through eating a healthful plant-based diet.

> Yeah, the whole reason why, I’m diabetic, I have high blood pressure, I have psoriasis, which is really bad, I have—they’ve told me that I’m in jeopardy of my kidneys going or something, and so the first thing I wanted to do was get off of every diabetic medication that they had…I wanted to control the high blood pressure. That seems to be really going haywire. And I wanted to do something with my psoriasis, because I break out every so often. So that was really my cue. (PBLM Participant)

Weight loss was also an important catalyst for participation. One participant noted the PBLM Program was appealing as a way of losing weight in order to prevent the development of multiple chronic diseases that affect their family members. And for another participant the PBLM Program was viewed as a more appealing treatment option than bariatric surgery, which they noted, was significantly more invasive.

### Program Implementation

#### Facilitators

Survey, interview, and focus group participants were asked what makes the PBLM Program successful. Three main themes repeatedly discussed included: 1) the PBLM team; 2) the approach used by the PBLM providers; and, 3) program resources (See Table 3).

**Table 3:** Facilitators and Challenges with the Implementation of the PBLM Program.

|  |  |
| --- | --- |
| **Facilitators** | **Challenges** |
| **Team composition**<br>The program team was comprised of providers with the right mix of credentials, expertise, and personalities to form a cohesive unit. Providers have personal experience of following a healthful plant-based diet. | **Administrative/system**<br>A lack of sufficient administrative resources within the hospital setting |
| **Approach**<br>Providers build rapport with participants, which yields a sense of accountability. The program is intentionally flexible and providers are non-judgmental. The program is individualized based on participants' needs. The program's focus is on a healthy lifestyle rather than weight loss. The program utilizes group classes that build a sense of community. | **Insurance/billing**<br>Confusion on the billing structure of the program and insurance coverage for services |
| **Resources**<br>Free resources serve to educate, inspire, and provide increased access to healthy foods for participants. | **Barriers to access/program capacity**<br>Program visits were only offered during workday hours and physician visits were restricted to one day per week, which were barriers for some participants. Program capacity was limited by large caseloads and a small number of program providers. |

#### The Team

A common cited facilitator was that the PBLM providers, as a unit, are key to the program’s value. Both program participants and PBLM providers described the importance of having the right mix of credentials, expertise, and personalities to form a cohesive unit.

> I think it’s really important to have all three components, because I found both [health coach name] and [registered dietitian name] helpful, and I learned. But I think most of all was what I get from when I meet with [physician name], but it wasn’t very often…but I think it’s so important to have all three: the doctor, the dietitian, and the lifestyle person. (PBLM Participant)

Each physician on the PBLM team not only has a deep knowledge of healthful plant-based nutrition, but also knowledge of their own specialty or area of focus in medicine. Multiple team members described the physicians’ role on the team as providing credibility to the healthful plant-based diet approach because participants strongly respect their physician’s expertise, and are thus are more likely to buy into the program and follow the recommendations.

> I think the first thing is that they have a professional medical doctor say to them, ‘This is what you need to do. This is proven. This will make you better.’ That is huge. That alone gives credibility. (PBLM Provider)

Multiple providers opined that participants respect their professional opinion more-so because they have personal experience following a healthful plant-based diet.

> Everybody will have their strengths and weaknesses, but I have to say, being plant-based for as long as I have definitely helps because we can really help people in different situations. And many of them ask me ‘Are you plant-based?’ I say ‘Yes, for 10 years, and I’ve been vegetarian for 17 before that,’ so that is huge because who’s going to listen to your advice if you’re not even doing it? (PBLM Provider)

Another PBLM provider talked about how sharing their own challenges with following a healthful plant-based diet can help their participants relate to them better and feel more comfortable.

> I think that as physicians, we’re kind of discouraged from sharing our personal experience with patients. But that said, every time I do share, I feel like it resonates with patients and is invaluable for them to hear when you make yourself a little vulnerable and you say, ‘Hey, I still crave chocolate bars. I’m still human.’ That really normalizes how patients are feeling. (PBLM Provider)

#### The Approach

Apart from the qualities of the PBLM providers themselves, their approach to working with participants was viewed as a key facilitator. The idea of building relationships between the PBLM providers and the participants was highlighted by both providers and participants. Participants described how they appreciate the providers making an effort to form connections with them on a personal level. PBLM providers similarly described the importance of getting to know the participants and building strong rapport with them from the start.

> I find that when I create a relatedness with a patient and they can really see that I’m interested in them, genuinely, my love for people is really what has made me successful in this because you create a safe space in this place, and people will start to really tell you everything, and they’ll open up to you. (PBLM Provider)

Interview respondents also explained how the relationship built between PBLM providers and participants led to a sense of accountability. Both the PBLM providers and participants talked about how working together places a sense of responsibility on the participant to follow-through and meet the health goals they have set together.

> They really feel like you’re invested in them, so I think they buy into the program more. I think they feel more held accountable, and they feel like somebody is working just as hard as they need to. So, I think that helps. (PBLM Provider)

While the PBLM Program creates accountability, respondents also stressed that the team uses an approach that is intentionally flexible and non-judgmental. Providers assessed where participants were in terms of their readiness to transition to a healthful plant-based diet and make other lifestyle changes, and worked with them at their own speed, whether that meant taking big leaps or baby steps, to make progress.

> … we approach patients in terms of meeting them where they are at and helping push them along but being non-judgmental around if they are not able to go…if they are going 50 percent plant-based, we don’t say ‘Oh, come on!’ But rather, ‘That’s great! That’s fantastic!’ It’s moving from Point A to Point B and respecting what the patient’s goals are and not rigidly saying, “Everyone has to go 100 percent plant-based in this program.’ Although that would be awesome, that’s just not what our approach is. (PBLM Provider)

Participants also commented on how they value the non-judgmental approach of PBLM providers, which is especially important for staying motivated after slips or deviations from the healthful plant-based diet.

> I saw [physician name] yesterday. She was very helpful because even though intellectually I understand that I’m doing better, I wasn’t really internalizing it. She said, ‘Think about the harm reduction. Even if you have a misstep, you’re still doing much better than you were and if you do well during the day, don’t give yourself such a hard time that it would knock you out of compliance at all.’ (PBLM Participant)

Similarly, the PBLM Program subscribes to an individualized approach, tailoring the intensity and duration of the program to meet individual needs. This approach has been viewed as a very positive aspect of the program, by both PBLM providers and participants.

> And so, there are some people that maybe need very infrequent one-on-one visits but more just group support just so they have that support system. Whereas other people— you can selectively maybe pick—this person needs weekly support because they’re having a hard time. And with that, you can probably better allocate resources. (PBLM Provider)

Another program facilitator mentioned by participants was the focus on living a healthy lifestyle rather than dieting or losing weight. This came up during focus groups when comparing the PBLM Program to more traditional weight loss programs.

> Because of Weight Watchers, I’m younger than you are, and I’ve gone since I was 15 with my mom. It’s a toxic culture… I’m trying to just be healthy and not focus on it as being a diet and restrictive. To me, I don’t respond well to that… (PBLM Participant)

When comparing the program to past experiences, participants talked about how the PBLM Program emphasized making healthy behaviors a part of one’s lifestyle, which in turn, improves health in a way that is more sustainable than temporary diets or weight loss programs.

> We’ve been talking about how do I make these things a habit that can be translated for anything. It doesn’t matter if I’m on vacation or I’m working full-time or if I’ve got to be at work at 9:00 a.m. now. I still have to keep the same habits. (PBLM Participant)

Another crucial aspect of the PBLM Program is the community formed through group classes and other social forums such as the private Facebook group. Both PBLM providers and participants agreed that these opportunities foster a sense of community, belonging, and social support that is important to their success.

> And they can still participate in the group if they like because the other thing is really that peer-learning piece. I really like to capitalize on that because I like people to hear from each other a lot. And me being a facilitator of that, that’s kind of my way with them, the way that I teach. (PBLM Provider)

An added bonus to the group classes is the ability for the program to engage more people, while also lessening the number of individual meetings participants need to have with each provider.

> Our hope was that with creating these classes, will also kind of lighten our load in that sense too, and accommodate more people. Because a couple of things were happening there: if we get working professionals, it’s really hard for people to take off time from their job to come in, and we’ve had people who are coming in from Long Island and all these really faraway places. And coming in every two weeks is just not feasible. (PBLM Provider)

#### Resources

The resources provided to all participants were viewed as another valuable aspect of the program. The registered dietitian and health coach described creating a starter guide for the program which acts as a master guide for following the healthful plant-based lifestyle, as well as a list of recommended books and websites. The guides also served to keep all program providers on the same page about specific nutritional recommendations. Program participants also referenced the cookbook they received from the program as a source of education and inspiration. A program provider described how providing participants with a cookbook that was free and approved by the program removed any confusion and indecision for participants in having to find the resource on their own. Finally, program participants reported utilizing the Healthy Savings Card and the HealthBucks they received from the program to save money on produce at local grocery stores and farmer’s markets. Both participants and providers viewed these as helpful tools that encouraged and increased access to healthy foods for participants.

#### Challenges

As expected with any new, pilot program, the PBLM Program encountered a range of challenges, some related to being part of a large healthcare system and others related to the program itself. Table 4 shows how participants responded to questions about difficulty with participation in the PBLM Program at the 6-month survey administration. Notably, the majority of participants (64.7%) reported difficulty reaching someone by phone, 41.8% of participants had difficulty rescheduling appointments, more than a third (34.7%) had difficulty scheduling appointments, and nearly 30% reported difficulty with getting insurance or billing questions answered. These were things that were generally outside of the program. Participants reported less difficulty with things that were in control of the program (i.e., interacting with clinic staff, completing the appointment).

**Table 4:** Difficulty with Participation in the PBLM Program at 6-Month Follow-up (n=84)

| **How easy or difficult were the following aspects of being involved with the program? (difficult/very difficult)** | **N** | **Percent** |
| --- | --- | --- |
| Starting the program (i.e., scheduling first appointment) | 74 | 24.3 |
| Reaching someone by phone | 68 | 64.7 |
| Making appointments | 72 | 34.7 |
| Rescheduling appointments | 67 | 41.8 |
| Getting insurance/Billing questions answered | 64 | 29.7 |
| Interacting with clinic staff | 71 | 12.7 |
| Completing all aspects of appointment (registering, vital signs, blood tests) | 71 | 19.7 |

NOTES: Ns may vary due to missing data.

**Table 5.** Program Satisfaction at 6-Month Follow-up (Agree/Strongly Agree) (n=84)

|  | **N** | **Percent** |
| --- | --- | --- |
| The information you were given about plant-based eating patterns was easy to understand. | 73 | 98.6 |
| The health coach had the appropriate knowledge and expertise to guide you. | 69 | 100.0 |
| You had enough time with or contact with the health coach to get your needs met. | 68 | 91.2 |
| The health coach was supportive. | 69 | 100.0 |
| Working with a health coach helped you. | 67 | 98.5 |
| The dietitian had the appropriate knowledge and expertise to guide you. | 69 | 95.7 |
| You had enough time with or contact with the dietitian to get your needs met. | 67 | 88.1 |
| The dietitian was supportive. | 69 | 98.6 |
| Working with a dietitian helped you. | 68 | 92.6 |
| The doctor had the appropriate knowledge and expertise to guide you. | 74 | 97.3 |
| You had enough time with or contact with the doctor. | 73 | 83.6 |
| The doctor you met with was supportive. | 74 | 95.9 |
| Working with the doctor helped you. | 72 | 90.3 |
| The program content will be useful to you in your life. | 73 | 98.6 |
| You are satisfied with your overall experience in the Plant-Based Lifestyle Medicine Program. | 71 | 85.9 |

NOTES: Ns may vary due to missing data.

Interview and focus group participants reported similar concerns to survey participants in more detail. The three primary areas of concerns were: (1) administrative/system-level challenges, (2) insurance/billing challenges, and (3) barriers to access and program capacity. Each of these challenges will be discussed in turn.

#### Administrative/System-level Challenges

Program leadership described difficulties associated with beginning an innovative pilot program within a traditional healthcare system. Namely, respondents cited a lack of sufficient administrative support. PBLM providers echoed the concerns shared by patients about contacting program staff and rescheduling appointments, as shown in Table 4. Below, a program provider described the difficulty associated with trying to navigate the hospital bureaucracy in order to launch additional components of the program.

> I’d say that there’s stuff that we want to do that is hard to do in a system that’s been designed for—like when we wanted to do a cooking class, and where do you have it, and the fire marshal, the equipment, all that stuff. But you know, the kinds of things that—the kinds of projects I want to see happen, or push forward, that’s probably been the biggest frustration, how things tend to move slowly in this kind of a system. There’s so many moving parts to everything that you want to get done. Sometimes your projects can kind of get lost in that, or it can take a long while, and then the excitement isn’t there as much as it was in the beginning. (PBLM Provider)

#### Insurance/Billing Challenges

Billing questions and variable insurance coverage for services proved to be a significant challenge for the program and its participants. This obstacle came up repeatedly during key informant interviews and also during the focus groups. Providers expressed enormous concern over the issue. At one point, the evaluation team was informed that enrollment of new patients and follow-up visits with the dietitian were paused temporarily to clarify issues related to commercial payor coverage of dietitian services. Program leadership described working with the hospital to resolve the situation, examining typical insurance coverage of dietitian services, as well as billing workflow changes related to the new electronic health system. In addition, there was a misconception among some patients that the program was free of charge, which led to surprise and frustration at insurance copayments or other out-of-pocket costs. Several respondents shared how insurance challenges impacted their ability to deliver or receive the program.

> I would hate not to continue my association, but I have to understand what I’m gonna be billed or not be billed for. So, that’s being ironed out. I hope they iron it out in my favor. I’ve been in the program about three months and right now, it’s on hiatus because I don’t know, every time I see a dietitian or I see the coach if I’m going to be billed. (PBLM Participant)

#### Barriers to Access and Program Capacity

While many people were initially very excited to participate in the PBLM Program, frequent visits became difficult for some participants due to the hours of operation and location of the program. These concerns were especially salient and became barriers to ongoing participation for those accessing the program from outer boroughs of New York City and beyond, and those with little autonomy over their daily schedules.

Limited capacity due to large caseloads and the need for repeat visits represented another very significant barrier to access. Without expanded hours, additional staff, or changes to the program, it became nearly impossible to ensure that there was fidelity to the program model of participants meeting with one of the program providers every two weeks.

> It feels like a lot. Now it feels like a lot. Earlier, we were able to get people every two weeks. If they wanted an appointment every two weeks, they got it. We were able to get new patients in within two weeks. Now, it’s a month, six weeks. Their revisit can’t be done until four weeks. That two-week appointment within the first six months is a big deal, so that might even be a nice thing. Dream—they see us each once for an initial, then they see us for revisits every two weeks for the first six weeks. (PBLM Provider)

### Program Satisfaction

Overall, satisfaction was extremely high among participants of the PBLM Program during the 6-month survey administration. Participants almost universally agreed or strongly agreed that the health coach, the dietitian, and the physicians had appropriate knowledge and expertise to help them. Similarly, the vast majority also said that the health coach, the dietitian, and the physicians were supportive. Again, nearly all agreed or strongly agreed that the program will be useful to them in their life. The one area for programmatic improvement may be offering more time with program providers and physicians. Even though satisfaction with the program was quite high, the three questions regarding having enough time with the program providers were the items that consistently had the lowest percentage of respondents who agreed or strongly agreed. That said, the vast majority of participants still thought they had enough time or contact with the health coach, dietitian, and physicians.

## DISCUSSION

This mixed-methods evaluation sought to assess the demand and feasibility of implementing the PBLM Program, which is, to our knowledge, the first lifestyle medicine program of its kind to be piloted within a traditional, safety-net healthcare setting. Our study provides important lessons for others seeking to implement similar programs in traditional clinical settings, and in particular, safety-net settings. Our findings show that demand for the program was extremely high and exceeded program capacity. Specifically, participants cited gaining more control over life, reducing medication, and weight loss as the main motivations for joining the program. Aspects of the program itself, including the program team, approach, and resources were successful and well-liked by participants. However, the program faced a lack of sufficient administrative support, and participants reported difficulties connecting with program and finance staff. Across all the interviewees - members of the PBLM team, participants, program dropouts - there was an unequivocal desire for the PBLM Program to continue. The program was said to be exceedingly valuable and unparalleled in the traditional medical care setting. Despite any challenges faced delivering the program or navigating participation, almost every person touched by the PBLM Program remained dedicated to its ongoing success. The recommendation from interviewees did not end with maintaining the program, there was a strong desire for it to expand in order to reach more individuals who would certainly benefit from participation.

Following the evaluation period, the PBLM Program was adapted and relaunched in March 2021 with several important modifications made in response to the findings from the pilot study. The eligibility criteria were narrowed by removing “overweight” and “high cholesterol” to allow limited resources to be focused on those with obesity and to discourage participation by individuals who were seeking lifestyle medicine as a sole therapeutic approach for familial hyperlipidemia. An electronic referral process was created to facilitate referrals by providers within the hospital system. The team also sought to more clearly set expectations for participation by creating a “Welcome Packet” with clear definitions of the program structure and duration, and answers to frequently asked questions, including information about insurance coverage. A program coordinator was hired to manage administrative issues and serve as a point of contact for patients. To improve program capacity and access, more emphasis was placed on group classes, which were expanded to evening hours and moved to an online-only format, and an additional physician was added to the team. Provider visits were converted to telephone and video visits in response to the COVID-19 pandemic, but also served to improve access to the program. Finally, the workflow was optimized for insurance coverage review and communication with patients around potential copayments or other out-of-pocket costs.

Although many important changes to the program and its operations were made, other models of care including shared medical appointments led by a physician along with a dietitian, health coach, and/or other staff are now being considered to address insurance coverage concerns for dietitian and health coach visits. Medicare and many commercial payors allow coverage of Medical Nutrition Therapy by dietitians only for diabetes, chronic kidney disease, and post-kidney transplant, and the number of billable hours per year is typically limited.[28] This poses a significant barrier for lifestyle programs operating in a traditional healthcare setting. The Medical Nutrition Therapy Act proposed in the U.S. Congress would expand the clinical conditions eligible for Medical Nutrition Therapy, but has not yet been passed. [28] On the other hand, shared medical appointments led by a physician, physician assistant, or an Advanced Provider Registered Nurse joined by a dietitian and/or health coach, are often covered by insurance and allow for more exposure to dietitian and health coach services that may not otherwise be reimbursable. [29]

The lessons learned from this pilot provide insight into important considerations for replication. Recommendations for others seeking to implement similar programs within traditional healthcare or safety-net settings include incorporating all the elements of lifestyle medicine, including diet, exercise, sleep, and stress management, to maximize efficacy of treatment; relying on a team approach comprised of trained providers with varying but complementary skill-sets; gaining buy-in from hospital administration to invest in a program; and finally, proactively clarifying billing structure and insurance coverage for services.

Two notable limitations related to the sample population affected this study. First, a combination of highly motivated people signing up for the program and few physician referrals unexpectedly led to a group of participants who were not necessarily representative of the typical safety-net hospital patient population, which is primarily comprised of racially and ethnically diverse individuals and those who have low socioeconomic status. Second, participants who consented to the research study (n=111) may have differed in demographics or other characteristics relative to the entire group of program participants (n=173).

Intensive plant-based lifestyle programs are proven to be an effective tool for reducing cardiometabolic risks, yet their availability is limited for underserved populations that may stand to benefit the most from participation. The PBLM Program pilot implementation aimed to bridge that gap, and was found to be exceedingly valuable by providers and participants alike, despite challenges faced delivering the program and navigating participation. Future research should focus on the sustainability of such programs in traditional clinical and safety-net settings, as well as rigorous randomized controlled trials of clinical outcomes as a result of program participation.

## Data Availability

All data produced in the present study are available upon reasonable request to the authors.

## ACKNOWLEDGMENTS

We thank Kayla Fennelly for helping schedule and conduct interviews, Sheetal Tolia for their help coding transcripts, Tanisha McIntosh for assisting with administrative functions related to the study, and all of the participants that took the time to talk to the study team about their experiences in the program.

## REFERENCES

1. Katz, D.L., et al., Lifestyle as Medicine: The Case for a True Health Initiative. Am J Health Promot, 2018. 32(6): p. 1452–1458.

2. Collaborators, U.S.B.o.D., et al., The State of US Health, 1990-2016: Burden of Diseases, Injuries, and Risk Factors Among US States. JAMA, 2018. 319(14): p. 1444–1472.

3. Ornish, D., et al., Intensive lifestyle changes for reversal of coronary heart disease. JAMA, 1998. 280(23): p. 2001–7.

4. Aldana, S.G., et al., The behavioral and clinical effects of therapeutic lifestyle change on middle-aged adults. Prev Chronic Dis, 2006. 3(1): p. A05.

5. Katz, D.L. and S. Meller, Can we say what diet is best for health? Annu Rev Public Health, 2014. 35: p. 83–103.

6. Kim, H., L.E. Caulfield, and C.M. Rebholz, Healthy Plant-Based Diets Are Associated with Lower Risk of All-Cause Mortality in US Adults. J Nutr, 2018. 148(4): p. 624–631.

7. Martinez-Gonzalez, M.A., et al., A provegetarian food pattern and reduction in total mortality in the Prevencion con Dieta Mediterranea (PREDIMED) study. Am J Clin Nutr, 2014. 100 Suppl 1: p. 320S–8S.

8. Satija, A., et al., Healthful and Unhealthful Plant-Based Diets and the Risk of Coronary Heart Disease in U.S. Adults. J Am Coll Cardiol, 2017. 70(4): p. 411–422.

9. Buckland, G., et al., Adherence to the Mediterranean diet and risk of coronary heart disease in the Spanish EPIC Cohort Study. Am J Epidemiol, 2009. 170(12): p. 1518–29.

10. Soltani, S., et al., Adherence to Dietary Approaches to Stop Hypertension Eating Plan and Prevalence of Irritable Bowel Syndrome in Adults. J Neurogastroenterol Motil, 2021. 27(1): p. 78–86.

11. Hauser, M.E., et al., Nutrition-An Evidence-Based, Practical Approach to Chronic Disease Prevention and Treatment. Supplement to the Journal of Family Practice, 2022. 71: p. S5–S16.

12. McMacken, M. and S. Shah, A plant-based diet for the prevention and treatment of type 2 diabetes. J Geriatr Cardiol, 2017. 14(5): p. 342–354.

13. Turner-McGrievy, G., T. Mandes, and A. Crimarco, A plant-based diet for overweight and obesity prevention and treatment. J Geriatr Cardiol, 2017. 14(5): p. 369–374.

14. Joshi, S., L. Ettinger, and S.E. Liebman, Plant-Based Diets and Hypertension. Am J Lifestyle Med, 2020. 14(4): p. 397–405.

15. Gupta, S.K., et al., Regression of coronary atherosclerosis through healthy lifestyle in coronary artery disease patients--Mount Abu Open Heart Trial. Indian Heart J, 2011. 63(5): p. 461–9.

16. de Lorgeril, M., et al., Mediterranean diet, traditional risk factors, and the rate of cardiovascular complications after myocardial infarction: final report of the Lyon Diet Heart Study. Circulation, 1999. 99(6): p. 779–85.

17. Benigas, S., D. Shurney, and R. Stout, Making the Case for Lifestyle Medicine. Supplement to the Journal of Family Practice, 2022. 71: p. S2–S4.

18. Englert, H.S., et al., The effect of a community-based coronary risk reduction: the Rockford CHIP. Prev Med, 2007. 44(6): p. 513–9.

19. Wright, N., et al., The BROAD study: A randomised controlled trial using a whole food plant-based diet in the community for obesity, ischaemic heart disease or diabetes. Nutr Diabetes, 2017. 7(3): p. e256.

20. American College of Lifestyle Medicine. Health Systems Council. 2021 February 8, 2022]; Available from: https://lifestylemedicine.org/ACLM/Partners/Health-Systems-Council/ACLM/Partners/Health-Systems/Health-Systems-Council.aspx?hkey=ca721dcc-8bfa-457c-b1cc-ee10e7866172.

21. GuidingStars. Heathy Savings. Available from: https://www.healthyrewards.com/.

22. NYC Department of Health and Mental Hygiene. Health Bucks. Available from: https://www1.nyc.gov/site/doh/health/health-topics/health-bucks.page.

23. Ortega, A.N., et al., Substantial improvements not seen in health behaviors following corner store conversions in two Latino food swamps. BMC Public Health, 2016. 16: p. 389.

24. American College of Lifestyle Medicine, L.L.U.H., Lifestyle Assessment Long Form Physician Version. 2017.

25. Development of the World Health Organization WHOQOL-BREF quality of life assessment. The WHOQOL Group. Psychol Med, 1998. 28(3): p. 551–8.

26. Lee, V., T. McKay, and C.I. Ardern, Awareness and perception of plant-based diets for the treatment and management of type 2 diabetes in a community education clinic: a pilot study. J Nutr Metab, 2015. 2015: p. 236234.

27. Ryan, R.M., R.W. Plant, and S. O’Malley, Initial motivations for alcohol treatment: relations with patient characteristics, treatment involvement, and dropout. Addict Behav, 1995. 20(3): p. 279–97.

28. Academy of Nutrition and Dietetics, Medical Nutrition Therapy Act. 2021. p. 1–7.

29. Lacagnina, S., et al., Lifestyle Medicine Shared Medical Appointments. Am J Lifestyle Med, 2021. 15(1): p. 23–27.

